# Risk of SARS-CoV-2 transmission by fomites: a clinical observational study in highly infectious COVID-19 patients

**DOI:** 10.1101/2022.03.22.22272773

**Authors:** Toni Luise Meister, Marielen Dreismeier, Elena Vidal Blanco, Yannick Brüggemann, Natalie Heinen, Günter Kampf, Daniel Todt, Huu Phuc Nguyen, Jörg Steinmann, Wolfgang Ekkehard Schmidt, Eike Steinmann, Daniel Robert Quast, Stephanie Pfaender

**Affiliations:** Department for Molecular & Medical Virology, Ruhr-University Bochum, 44801 Bochum, Germany; Department of Medicine I, St. Josef-Hospital Bochum, Ruhr-University Bochum, 44791 Bochum, Germany; Institute for Hygiene and Environmental Medicine, University Medicine Greifswald, 17475 Greifswald, Germany; European Virus Bioinformatics Center (EVBC), 07743 Jena, Germany; Department of Human Genetics, Ruhr-University Bochum, 44791 Bochum, Germany; Institute of Clinical Hygiene, Medical Microbiology and Infectiology, Paracelsus Medical University, Nuremberg, Germany

**Keywords:** Coronavirus, SARS-CoV-2, COVID-19, fomites, surfaces, cough, transmission, environmental stability

## Abstract

**Background:** The contribution of droplet-contaminated surfaces for virus transmission has been discussed controversially in the context of the current Severe Acute Respiratory Syndrome Coronavirus-2 (SARS-CoV-2) pandemic. Importantly, the risk of fomite-based transmission has not been systematically addressed.

**Methods:** We initiated this single-center observational study to evaluate whether hospitalized COVID-19 patients can contaminate stainless steel carriers by coughing or intensive moistening with saliva and to assess the risk of SARS-CoV-2 transmission upon detection of viral loads and infectious virus in cell culture. Fifteen hospitalized patients with a high baseline viral load (CT value ≤ 25) shortly after admission were included. We documented clinical and laboratory parameters and used patient samples to perform virus culture, quantitative PCR and virus sequencing.

**Results:** Nasopharyngeal and oropharyngeal swabs of all patients were positive for viral RNA on the day of the study. Infectious SARS-CoV-2 could be isolated from 6 patient swabs (46.2 %). While after coughing, no infectious virus could be recovered, intensive moistening with saliva resulted in successful viral recovery from steel carriers of 5 patients (38.5 %).

**Conclusions:** Transmission of infectious SARS-CoV-2 via fomites is possible upon extensive moistening, but unlikely to occur in real-life scenarios and from droplet-contaminated fomites.

## Background

The emergence of Severe Acute Respiratory Syndrome Coronavirus-2 (SARS-CoV-2), the cause of the Coronavirus Disease-19 (COVID-19), has raised the general awareness towards different hygiene and prevention measures to limit viral spread. Although SARS-CoV-2 is mainly transmitted via respiratory droplets and aerosols exhaled from infected individuals (e.g., upon breathing, speaking, coughing or sneezing [1]), droplet-contaminated surfaces (fomites) have also been widely perceived as another potential route of transmission. In particular, different studies reported that SARS-CoV-2 can persist on inanimate surfaces for days under controlled laboratory conditions [2–4] and genomic material of SARS-CoV-2 has been detected on diverse surfaces and materials in hospital, private and public settings [5]. Consequently, a clinically significant risk of SARS-CoV-2 transmission by fomites has been assumed and extensive hand hygiene and disinfection procedures have been initiated early during the pandemic worldwide. Although recent studies suggest a low risk of viral transmission by fomites for most instances [1,6], it is still considered possible given a timely order of events (e.g., direct contamination of a surface by an infected individual followed by timely skin contact by another individual and direct contact towards susceptible mucosae) [7]. However, most efforts to study surface transmission of SARS-CoV-2 have either focused on the detection of viral RNA via RT-qPCR rather than direct detection of infectious viral particles and/or employed lab-grown viruses which do not recapitulate the specific infectivity of patient-derived SARS-CoV-2 particles. Consequently, these findings do not necessarily allow to adequately estimate the potential of SARS-CoV-2 transmission from directly contaminated surfaces.

To examine the risk of transmission by surfaces directly after contamination by individuals infected with SARS-CoV-2, we performed a clinical observational study, including hospitalized patients with high viral loads (CT ≤ 25, up to 2.03×10^9^ RNA copies). The aim of this study was to evaluate if confirmed hospitalized COVID-19 patients can contaminate stainless steel carriers by coughing or intensive moistening with saliva and to assess the risk of SARS-CoV-2 transmission upon detection of viral loads and infectious virus in cell culture.

## Methods

### Study cohort

Hospitalized patients (age > 18 years) treated at St. Josef-Hospital Bochum, Germany, with confirmed SARS-CoV-2 infection with a high virus load (RT-PCR from combined nasopharyngeal and oropharyngeal swab (swab) with a cycle threshold (CT) ≤ 25 on admission) were included in this study. The initial viral load was determined using Allplex™ 2019-nCoV Assay (Seegene Inc., Seoul, Republic of Korea) targeting three SARS-CoV-2 specific genes (E gene, RdRP gene and N gene) with a sensitivity of 100 copies per run. Exclusion criteria were acute myocardial infarction, current need of ventilation support systems (e.g., high flow-or non-invasive ventilation), current treatment in an intensive care unit, evidence of drug or alcohol abuse, acute psychiatric disorders and any clinical or mental disorder that might deteriorate the patient’s condition during the standardized procedure of sampling (such as dysphagia), as per investigator’s judgement. Collected clinical data included medical history, current daily medication, laboratory results, blood gas analysis, and results of x-rays or computed tomography (to define the “clinical classification of COVID-19-infection” following the recommendation of the World Health Organization (WHO) [8] and Robert Koch Institute (RKI) [9]).

### Study design

After written informed consent, two combined nasopharyngeal and oropharyngeal swabs were collected from each patient. Then, patients were asked to forcefully cough two-times on a pre-defined surface area containing nine standardized steel-carriers, each with a one-centimeter diameter (“cough”), using a specially designed tripod with a defined distance of 15 centimeters (Appendix Figure 1). In addition, patients were asked to moisten nine steel carriers with saliva for ten seconds within their mouth (“moisten”). After defined time points at room temperature (1 min, 5 min, 15 min, 30 min, 45 min, 90 min, 120 min and 240 min), the steel-carriers were placed in containers containing 2 mL cold Dulbecco’s modified Eagle’s medium (DMEM complete, supplemented with 10 % (v/v) fetal calf serum, 1 % non-essential amino acids, 100 IU/mL penicillin, 100 μg/mL streptomycin and 2 mM L-Glutamine) and transported on ice to the biosafety level three laboratory of the Ruhr-University Bochum. The study was conducted between November 2020 and April 2021.

### Virus culture

VeroE6 cells were seeded at 3×10^5^ cells/well in a six well cell culture plate and incubated for at least four hours at 37 °C and 5 % CO_2_. Hereafter, the medium was replaced with 1·8 mL of patient swabs, “cough” samples or “moisten” samples and 2·5 μg/mL amphotericin B was added. Over a maximum period of ten days, cells were monitored daily for the appearance of a cytopathic effect (CPE), indicating productive virus infection. Upon visible CPE, cells were harvested for RT-qPCR and the supernatant (SN) was collected for viral titration and RT-qPCR. Viral titers in the SN were quantified by endpoint-dilution and the 50 % tissue culture infective dose (TCID50/mL), calculated according to Spearman and Kärber [10].

### Reverse Transcription Quantitative PCR (RT-qPCR)

SARS-CoV-2 RNA was isolated from the supernatant using AVL buffer and the QIAamp Viral RNA Kit (QIAGEN®, www.qiagen.com) according to the manufacturer’s instructions. RNA was directly subjected to one-step quantitative PCR (RT-qPCR) running a GoTaq Probe 1-Step RT-qPCR System (Promega®, www.promega.com). Total RNA was purified from VeroE6 cells using the RNeasy Mini Kit (QIAGEN®, www.qiagen.com). Subsequently, 500 ng of total RNA were reverse transcribed using the PrimeScript™ RT Master Mix (Takara®, www.takarabio.com) and subjected to two-step RT-qPCR running a GoTaq Probe 2-Step RT-qPCR System (Promega®, www.promega.com). RT-qPCR was performed as described previously [11] using a light cycler LC480 to quantify the M-Gene abundance.

### Data analysis and sample size

Clinical patient parameters are expressed as mean ± SD or n (% of total). Results are expressed as means (± SEM). Clinical characteristics were screened for correlations using Spearman’s correlation coefficient. Statistical significance was defined as α=0·05. Statistical analysis was performed using GraphPad Prism version 8.0.0 for Windows (GraphPad Software, San Diego, California USA, www.graphpad.com). Sample size calculation was performed using G*Power Version 3.1.9.6 for windows [12].

### Legal and ethical considerations

The study was conducted according to the revised principles of the Declaration of Helsinki and was approved by the ethics committee of the Ruhr-University Bochum (registration number 20-7065) in November 2020. All patients gave written informed consent.

### Sequencing and strain assignment

RNA of the initial swaps was isolated using the NucleoSpin RNA kit (Macherey & Nagel) followed by a reverse transcription utilizing the SuperScript IV together with Oligo dT and random hexamer primer (Thermo Fisher) according to the manufacturers’ instructions. Subsequently, the cDNA was subjected to deep sequencing. Sequencing libraries were prepared from 4·5μl cDNA using NEBNext® ARTIC SARS-CoV-2 Library Prep Kit for Illumina sequencing platforms (New England BioLabs® Inc., neb.com, catalog #E7650). Concentration and size of the cDNA amplicons and libraries were assessed using Qubit fluorometer and Tapestation (High Sensitivity D1000 ScreenTape), respectively. High throughput paired end sequencing was performed using Illumina MiSeq sequencer and MiSeq Reagent Kit v2 (500-cycles) following the manufacturer’s recommendations. Raw reads were quality checked, trimmed and mapped to the SARS-CoV-2 reference sequence (NCBI Reference Sequence: NC_045512) using QIAGEN CLC Genomics Workbench 21.0.5. After removing duplicates, partially full length consensus sequences were extracted and samples were assigned to respective lineages using the pangolin tool [13] (Table 2). In addition, variants in spike domains were identified and annotated using Geneious prime 2021.2.2 (https://www.geneious.com).

## Results

### Study cohort

A total of 15 patients (33.3 % female) between 39 and 89 years (mean age 70·5 (± 12·5) years) were recruited. Baseline parameters including laboratory findings on admission are presented in Table 1. All study patients had risk factors for a severe course of COVID-19 according to the criteria of the WHO [8] and RKI [14]. Three (20 %) patients had up to two risk factors, while most patients had multiple risk factors for a severe COVID-19 illness (3-4 risk factors: 7 (46·7 %), 5-6 risk factors: 2 (13·3 %), >6 risk factors: 1 (6·7 %)). Body temperature on admission was 37·0 (± -0.9) °C. On the study day, 9 (60 %) patients were categorized with a mild COVID-19 disease according to STACOB criteria (adaptation following the WHO Therapeutics and COVID-19:living guideline [8]). Nasal oxygen support was required by seven (46·7 %) patients. Mean peripheral oxygen saturation on admission was 93·4 % (SEM ± 7·4 %). None of the included patients were vaccinated against SARS-CoV-2.

**Table 1:** Baseline parameters including laboratory findings of the study group.

| Parameter | Unit | Normal range | Result |
| --- | --- | --- | --- |
| Participants | n | n/a | 15 |
| Age | years | n/a | 70.5 (± 12.3) |
| Male / female (% female) | n | n/a | 10/5 (33.3 %) |
| BMI | kg/m<sup>2</sup> |  | 28.8 (± 6.0) |
| Arterial hypertension | n | n/a | 10 (66.7 %) |
| - ACE inhibitors | n | n/a | 3 (20 %) |
| Dyslipidemia | n | n/a | 5 (33.3 %) |
| Diabetes mellitus | n | n/a | 5 (33.3 %) |
| Currently smoking | n | n/a | 8 (53.3 %) |
| Family history of CVD | n | n/a | 4 (26.7 %) |
| Immunosuppression | n | n/a | 5 (33.3 %) |
| Active malignancy | n | n/a | 4 (26.7 %) |
| Chronic pulmonary disease | n | n/a | 7 (46.7 %) |
| Chronic renal disease | n | n/a | 3 (20 %) |
| Leucocytes | /μl | 4600-9500 | 6182.7 (± 3560.0) |
| Hemoglobin | g/dl | 14.0-18.0 | 11.5 (± 2.6) |
| Thrombocytes | /μl | 150000-400000 | 175933.3 (± 64077.5) |
| D-dimers | μg/ml | <0.5 | 1.3 (± 0.9) |
| Lactate dehydrogenase | U/l | 135-225 | 250 (± 63) |
| eGFR | ml/min/1.73m<sup>2</sup> | >90 | 63.4 (± 26.4) |
| Procalcitonin | ng/ml | <0.5 | 33.5 (± 124.9) |
| C-reactive protein | mg/l | <5.0 | 41.8 (± 38.9) |
| pH |  | 7.35-7.45 | 7.4 (± 0.1) |
| Bicarbonate | mmol/l | 22-26 | 26.1 (± 4.3) |
| Peripheral oxygen saturation | % | ≥95 | 93.4 (± 7.4) |
| Air Temperature | °C | n/a | 5.5 (± 5.5) |
| Air humidity | % | n/a | 72.8 (± 18.7) |

Data are presented as n (% of total) or mean (± SD). Risk factors for severe COVID-19 were defined as age > 50 years, male sex, smokers, adiposity (BMI >30), Downs syndrome, history of cardiovascular diseases, chronic lung disease, chronic renal disease, psychiatric diseases, diabetes mellitus, malignant or hemic diseases or immunodeficiency [14]. eGFR was estimated using Chronic Kidney Disease Epidemiology Collaboration (CKD-EPI) equation [27]. Peripheral oxygen saturation was determined on a finger, using a pulse oximeter. ACE = angiotensin converting enzyme. BMI = body mass index. CVD = cardiovascular disease. eGFR = estimated glomerular filtration rate.

On the day of the study, most patients had only mild symptoms (n= 9/15, 60 %) and were categorized to mild COVID-19 disease. Follow-up revealed a clinical worsening in 10 patients and 3 died (2 patients died of COVID-19, 1 patient died with COVID-19). Consistent with literature [15], a high level of lactate dehydrogenase (r=0·53, p=0·044), leucocytes (r=0·69, p=0·0056), but also C-reactive protein (r=0·54, p=0·035) on admission was significantly correlated with a more severe COVID-19 infection (Appendix Figure 2). No significant correlation was found for COVID-19 severeness and other described severeness predictors including age (r=0·44, p=0·11), alanine aminotransferase (r=-0·31, p=0·26), aspartate aminotransferase (r=0·07, p=0·81), procalcitonin (r=0·53, p=0·47), or D-dimers (r=0·45, p=0·11).

### SARS-CoV-2 viral load and sequencing

The patients’ viral load before sample acquisition as determined with the combined nasopharyngeal and oropharyngeal swabs are displayed in Table 2. Mean CT-values in RT-qPCR analyses were E-Gene 15.3 (SEM ± 2·7), S-Gene 18·1 (SEM ± 4·9) RdRP-Gene 17·2 (SEM ± 3·7) and N-Gene 19·4 (SEM ± 5·5). Viral variants included supposed wildtype (n=10, 66·7 %), variant of concern (VoC) Alpha (n=4, 26·7 %) and VoC Beta (n=1, 6·7 %). Deep sequencing using the pangolin tool [13] confirmed lineage assignment for 14/15 (93·3 %) samples for nearly full-length genomes. Variant patterns identified by detailed investigation of spike domains underlined lineage assignment according to RKI variant reports [16] (Figure 1, Table 2).

**Table 2:** Viral load before sample acquisition using Naso Oropharyngeal swabs.

| **Patient** | **Initial CT values**<br>(*E-Gene/S-Gene/RdRP-Gene/N-Gene; VOC-Analyses\**) | **Deep Sequencing** | **Mutations identified in spike domains** |
| --- | --- | --- | --- |
| P1 | 13/ - /17/15 | B.1.389 | **D614G**, T723I |
| P2 | 14/ - /16/17 | B.1.1.70 | **D614G** |
| P3 | 13/ - /14/15 | B.1.1.70 | **D614G** |
| P4 | 16/ - /17/20 | B.1.1.70 | **D614G** |
| P5 | 12/ - /13/14 | B.1.221 | S98F, **D614G** |
| P6 | 17/ - /18/16 | B.1.177.7 | A222V, **D614G** |
| P7 | 21/ - /22/19 | B.1.177 | A222V, **D614G**, A1020V |
| P8 | 16/ - /16/18 | B.1.221 | S98F, L141LF, **D614G** |
| P9 | - /16/16/14 | B.1.177 | A222V, **D614G** |
| P10 | - /12/13/13 | B.1.1.153 | **D614G** |
| P11 | - /24/25/23; VOC B1.351* | B.1.351 | H69Y, **D80A**, **D215G**, del242-244, **K417KT**, **E484EK**, **N501NY**, **D614G**, A701V |
| P12 | - /24/23/32; VOC B1.1.7* | - # | - # |
| P13 | - /17/16/25; VOC B1.1.7* | B.1.1.7 | **del144**, **N501NY**, **A570D**, **D614G**, **P681H**, A694AS, **T716I**, **S982A**, **D1118H** |
| P14 | - /13/12/21; VOC B1.1.7* | B.1.1.7 | **del69-70**, S98F, D138DH, H245Y, **N501NY**, **A570D**, **D614G**, **P681H**, A694AS, **T716I**, **S982A**, **D1118DH** |
| P15 | - /21/20/29; VOC B1.1.7* | B.1.1.7 | **del69-70**, **del144**, **N501Y**, **A570D**, **D614G**, **P681H**, A694AS, **T716I**, **S982A**, **D1118H** |

Data is presented as individual data. Lineage of deep sequencing is presented according to the pangolin tool [13]. CT = cycle threshold resulting from RT-PCR performed at St. Josef-Hospital Bochum. VoC = variants of concern. \*VoC-Analyses started in the midst of the study period in February 2020 and was performed via melting curve analysis.
#no sufficient material available. **bold:** characteristic mutations associated with VOC according to RKI [16].

**Figure 1:**
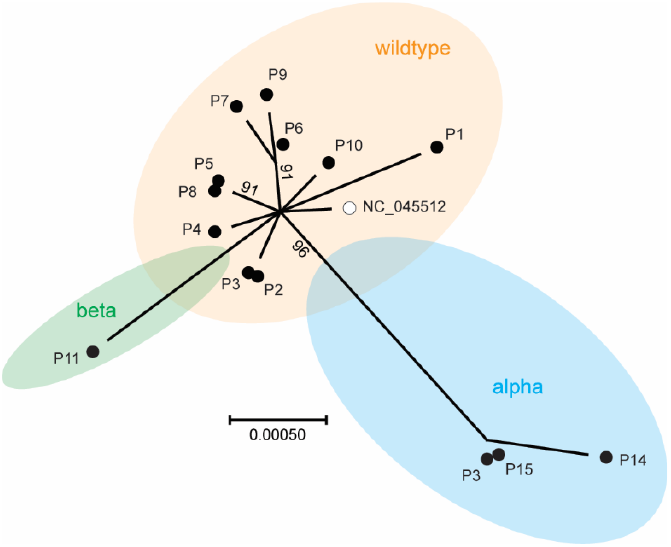
Viral Isolates analyzed in this study. Spike domains (nt) of 14 samples (black dots) and the SARS-CoV-2 reference genome NC_045512 (open circle) were assembled using Clustal Omega at EMBL-EBI [28]. Phylogenetic analysis was conducted with “One click” at Phylogeny.fr [29]and tree was visualized with MEGA X [30]. Scale bar indicates the number of changes per site in maximum likelihood inference (HKY85 substitution model); numbers at branches represent bootstrap values (1000 repetitions; cut-off ≥ 70%).

Two patient samples were excluded from the study due to bacterial/fungal contamination within the cultures (Appendix Table 1). Viral RNA could be detected from all (n=13/13) combined nasopharyngeal and oropharyngeal swabs (inoculum) and viral RNA could be successfully detected within the inoculated cell cultures (Figure 2, Appendix Figure 3). After inoculation with the “swabs”, viral loads in the cells ranged from 2·23×10^1^ to 2·03×10^9^ RNA copies/50 ng and in the supernatant from not detectable to 6·58×10^7^ RNA copies/mL (Appendix Table 1). Infectious virus (“Infectivity”), determined as TCID_50_/mL, could be recovered from the nasal-oropharyngeal swabs from n=6/13 (46·2 %) patients (Figure 2; P1, P2, P3, P5, P8, P10). Of note, despite inclusion criteria defining a high viral load (CT ≤ 25, Table 2), some patients displayed lower viral loads at the time-point of the study, with none of the swab samples resulting in productive virus infection in cell culture (Appendix Figure 3; P.11, P12, P13, P14).

**Figure 2:**
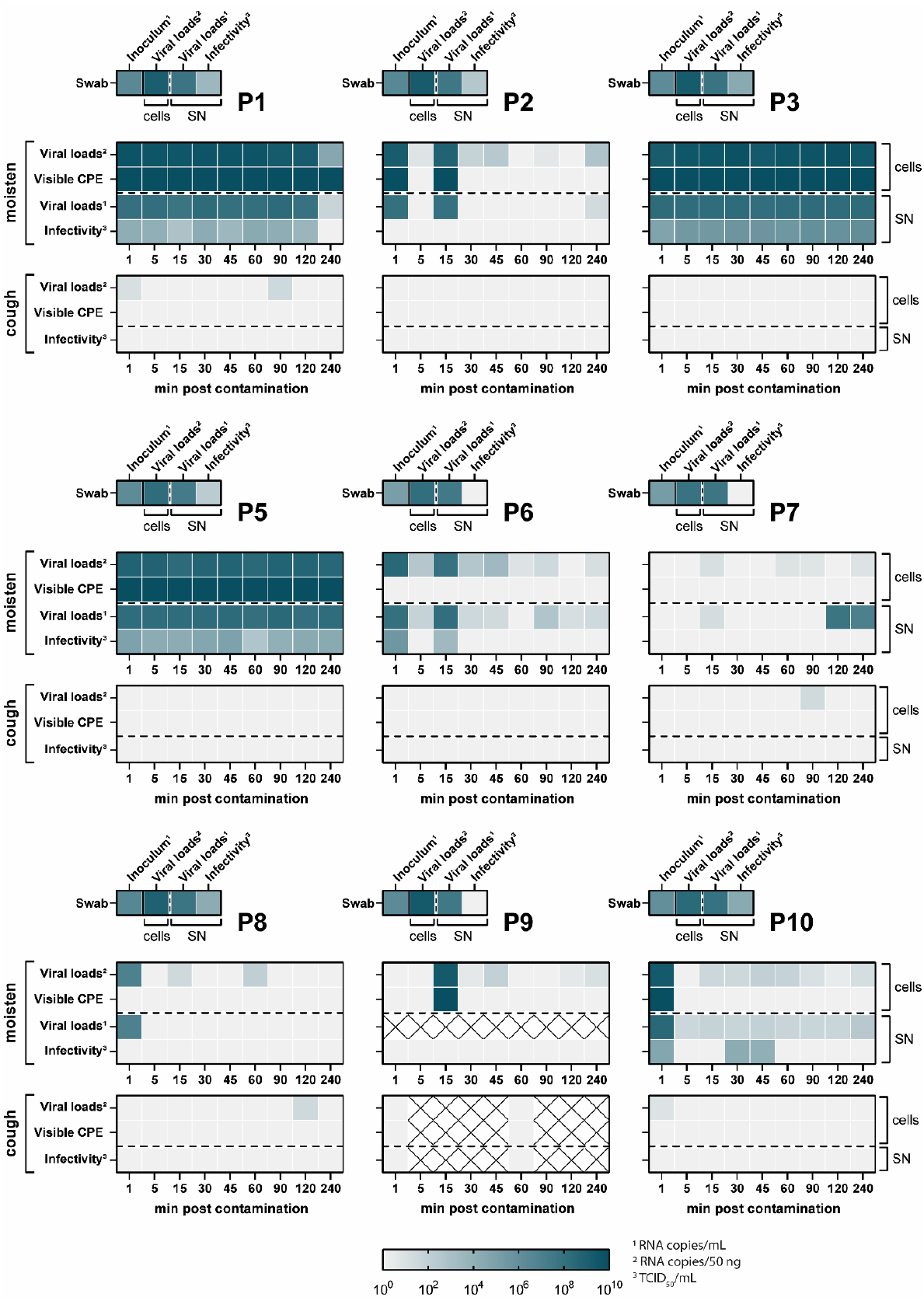
Quantification of viral loads and infectivity of patient swabs, “moisten” samples and “cough” samples that could be successfully recovered in cell culture. VeroE6 cells were inoculated with the patient material and monitored on a daily basis. Upon the emergence of cytopathic effects, the supernatant was collected to determine viral loads by RT-qPCR (RNA copies/mL; indicated by ^1^) and viral titers by an endpoint-dilution assay (TCID_50_/mL, indicated by ^3^). Additionally, RNA was isolated from the cells and subjected to RT-qPCR to determine viral loads (RNA copies/50 ng total RNA, indicated by ^2^). For each patient (P1-P10), three panels were designed. The top small panel includes exclusively data regarding the patient swabs, while the larger middle panel shows the data for the “moisten” samples and the lower panel the data collected from the “cough” samples. For “moisten” and “cough” samples viral loads and infectivity at nine different time points were determined. The color indicates the amount of virus being detectable in each sample, with light grey being the lower limit of detection to dark blue resulting in 10^10^ RNA copies/mL, RNA copies/50 ng or TCID_50_/mL. The visible CPE was rated two dimensionally, with light grey being “no visible CPE” and dark green being “visible CPE”.

### Evaluation of SARS-CoV-2 transmission risk

Steel-carriers contaminated via intensive moistening with saliva (Figure 2, “moisten”) resulted in a visible CPE and detectable viral RNA within the cells (at least one time point positive) in n=6/13 (46·2 %) cases (P1, P2, P3, P5, P9, P10). Despite the absence of a visible CPE in patient 6 and patient 10 (Figure 2, time points 30 and 45 min), viral RNA was detected in the cells and supernatants, and infectious virus was quantified, indicating that harvesting might have been too early for the appearance of CPE. Infectious virus was recovered from n=5/13 (38·5 %) contaminated steel carriers (Figure 2; “SN infectivity” P1, P3, P5, P6, P10) with viral titers ranging from 5·59×10^1^ to 8·68×10^5^ TCID_50_/mL. In some patients, infectious virus could be recovered from the steel-carriers for up to 240 min after incubation at room temperature (Figure 2; P1, P3, P5), underlining the environmental stability of SARS-CoV-2 over several hours. For other samples (Figure 2; P2, P6, P10), infectious virus could be recovered at early time points only (1 min, 15 min, 30 min, 45 min).

After contamination with coughing, viral RNA could be weakly detected within five of the cellular samples (Figure 2; “cough” P1, P7, P8, P10, Appendix Figure 3; P11). However, none of the via coughing contaminated surfaces resulted in productive cellular infection as determined by the appearance of CPE and quantification of infectivity (Figure 2, Appendix Figure 3).

## Discussion

While respiratory droplets and aerosols exhaled from infected individuals are currently considered the main route of SARS-CoV-2 transmission, the role of droplet-contaminated surfaces (fomites) as a potential source of infection remains controversial. Fomite-based transmission has been proposed to contribute to the spread of other common respiratory pathogens [17,18], including experimental studies examining the transfer of infectious influenza viruses and/or respiratory syncytial virus between hands and surfaces [19]. However, current evidence points towards a low risk of SARS-CoV-2 transmission in this scenario [1,20], requiring a timely order of specific events [7]. To examine this potential risk of SARS-CoV-2 surface transmission, we assessed the amount of SARS-CoV-2 genomic material and infectious viral particles after contamination by individuals infected with SARS-CoV-2 over time. The results of the present study highlight that viral contamination via coughing on surfaces does not represent a major risk of transmission.

Our study cohort was characterized by non-vaccinated, hospitalized, mostly elderly patients with multiple comorbidities. Since the vaccination program in Germany started in December 2020 (during the study period), none of the patients were vaccinated against SARS-CoV-2. This cohort may therefore be quite representative for hospitalized patients during the first and second COVID-19 wave in most countries [21]. However, since the proportion of unvaccinated people in the population remains significant [22] and completely vaccinated individuals can still be infected and often present with high viral loads [23], the present results remain of significant importance for the ongoing pandemic. Initially, we were able to recruit patients in early stages of COVID-19 due to outbreaks in hospitals and rehabilitation facilities and subsequently early referral to the isolation ward. Later, patients were diagnosed in ambulatory settings and predominantly admitted via the emergency department, often only when clinical conditions deteriorated and ambulatory management failed. The hospitalized cohort may explain the high rate of risk factors and, consequently, follow-up mortality in the present study. However, the present cohort is characterized by high viral load. Hence, a high transmission rate can be assumed [24], supporting the main conclusion of the study. Of note, several laboratory parameters on admission significantly correlated with a severe outcome. However, the present study was not powered for this analysis and therefore, these correlations need to be considered exploratory.

Infectious virus could be recovered from the combined nasopharyngeal and oropharyngeal swabs and steel-carriers contaminated via intensive “moistening” from a significant number of patients. For some patients, infectious virus could be recovered for up to 240 min (Figure 2). This demonstrates that infectious virus can be transferred from saliva by moistening onto surfaces from patients and can be recovered for several hours. As described previously, we did not observe differences of the viral stability between the wildtype and VoCs (Alpha and Beta), implying a comparable environmental stability [25]. The stability of SARS-CoV-2 on surfaces is likely determined by a combination of factors, including the initial amount of infectious virus deposited, possible presence of antibodies within the sputum and environmental parameters. Given the controlled laboratory conditions for virus recovery as herein presented (e.g., large inoculums, small surface area, no UV exposure), the viral survival observed might therefore differ from real-life scenarios, necessitating careful interpretation. For example, a recent study observed a low transfer efficiency between different surfaces and fingertips following an initial drying of an inoculum with a low viral titer (1×10^4^ TCID_50_/mL) [20]. Hence, even if sufficient viable virus is deposited on a surface, a timely contact and high transfer efficiency are required to transfer an infectious dose, which subsequently needs to be exposed towards susceptible tissues (e.g., mucosa, eyes). Importantly, we did not observe the recovery of infectious virus after patients coughed onto a surface, implying that droplet-contamination of surfaces does not present a major transmission route for SARS-CoV-2. Given that non-hospitalized and pre-symptomatic, asymptomatic, and mildly symptomatic individuals across different age groups frequently display viral loads within a comparable range [26], similar observations as herein observed for elderly and hospitalized patients can be inferred.

Our study encompasses several limitations. Patients were encouraged to forcefully cough twice to contaminate surfaces. However, we cannot exclude that potentially repeated coughing over a prolonged time results in a more effective virus transfer compared to our controlled conditions. Moreover, sneezing can produce significantly more infectious droplets potentially containing infectious particles, therefore, we cannot exclude potential transmissions via other this route. Furthermore, a selection bias cannot be excluded, and the included patients are not demographically representative. Strengths of the present study include the high viral load of the patients included, a standardized protocol for sample acquisition, laboratory procedures and the inclusion of VoC.

## Conclusion

The present study provides evidence that fomites may not be as critical in the transmission of SARS-CoV-2 as initially suspected. However, the present study also provides evidence that infectious SARS-CoV-2 can be found on some fomites after contamination with extensive amounts of saliva. Therefore, common hygiene practices (e.g., coughing/sneezing into elbows, hand hygiene) should still be considered to avoid the surface contamination and virus transfer. Face masks may further mitigate the risk of fomite transmission. Collectively, our findings suggest that fomites contaminated with coughing are unlikely to be an important source of SARS-CoV-2 transmission.

## Data Availability

All data produced in the present work are contained in the manuscript

## Declarations

### Consent for publication

The study is published with patients consent.

### Availability of data and materials

The datasets used and/or analyzed during the current study are available from the corresponding author on reasonable request.

### Author Contributions

Conceptualization, T.L.M., M.D., W.E.S., E.S., D.R.Q., S.P.

Methodology, T.L.M., M.D., D.R.Q., S.P.

Investigation, T.L.M., M.D., E.V., N.H.

Writing –Original Draft, S.P., Y.B.

Writing –Review & Editing, T.L.M., M.D., E.V., N.H., G.K., D.T., H.P.N., J.S., W.E.S., E.S., D.R.Q., S.P.

Visualization, T.L.M., D.T., S.P.

Resources, W.E.S., E.S., D.R.Q., S.P.

Supervision, D.R.Q., S.P.

## Acknowledgements

We would like to thank all members of the Molecular and Medical Virology at the Ruhr-University Bochum for their support and fruitful discussions. Furthermore, we are grateful for the support of the healthcare workers at St. Josef-Hospital Bochum and appreciate all patients who volunteered to participate in this study.

