## Supplementary Material for "Risk of SARS-CoV-2 transmission by fomites: a clinical observational study in highly infectious COVID-19 patients"

***Short title: Risk of SARS-CoV-2 transmission by fomites***

Toni Luise Meister, M.Sc. [1*], Marielen Dreismeier, M.D. [2*], Elena Vidal Blanco [1], Yannick Brüggemann, Dr. [1], Natalie Heinen, M.Sc. [1], Günter Kampf, Prof. [3], Daniel Todt, Dr. [1,4], Huu Phuc Nguyen, Prof [5], Jörg Steinmann, Prof. [6], Wolfgang Ekkehard Schmidt, Prof. [2], Eike Steinmann, Prof. [1], Daniel Robert Quast, Dr. [2#] and Stephanie Pfaender, Prof. [1#]

[1] Department for Molecular & Medical Virology, Ruhr-University Bochum, 44801 Bochum, Germany

[2] Department of Medicine I, St. Josef-Hospital Bochum, Ruhr-University Bochum, 44791 Bochum, Germany

[3] Institute for Hygiene and Environmental Medicine, University Medicine Greifswald, 17475 Greifswald, Germany

[4] European Virus Bioinformatics Center (EVBC), 07743 Jena, Germany

[5] Department of Human Genetics, Ruhr-University Bochum, 44791 Bochum, Germany

[6] Institute of Clinical Hygiene, Medical Microbiology and Infectiology, Paracelsus Medical University, Nuremberg, Germany

* These authors contributed equally and share first authorship.

[#] These authors contributed equally and share last authorship.

**Abstract**

Background:

The contribution of droplet-contaminated surfaces for virus transmission has been discussed controversially in the context of the current Severe Acute Respiratory Syndrome Coronavirus-2 (SARS-CoV-2) pandemic. Importantly, the risk of fomite-based transmission has not been systematically addressed.

Methods:

We initiated this single-center observational study to evaluate whether hospitalized COVID-19 patients can contaminate stainless steel carriers by coughing or intensive moistening with saliva and to assess the risk of SARS-CoV-2 transmission upon detection of viral loads and infectious virus in cell culture. Fifteen hospitalized patients with a high baseline viral load (CT value ≤ 25) shortly after admission were included. We documented clinical and laboratory parameters and used patient samples to perform virus culture, quantitative PCR and virus sequencing.

Results:

Nasopharyngeal and oropharyngeal swabs of all patients were positive for viral RNA on the day of the study. Infectious SARS-CoV-2 could be isolated from 6 patient swabs (46.2 %). While after coughing, no infectious virus could be recovered, intensive moistening with saliva resulted in successful viral recovery from steel carriers of 5 patients (38.5 %).

Conclusions:

Transmission of infectious SARS-CoV-2 via fomites is possible upon extensive moistening, but unlikely to occur in real-life scenarios and from droplet-contaminated fomites.

**Appendix Table 1: Viral loads and viral titers of samples obtained after cultivation on cell cultures**

| Patient | Sample Type | cells | | supernatant | |
|---|---|---|---|---|---|
| | | Viral loads (RNA copies/50 ng) | Visible CPE | Viral loads (RNA copies/mL) | Infectivity ($TCID_{50}$/mL) |
| P1 | Swab | $1.10\times10^{9}$ | 4 d p.i. | $4.34\times10^{7}$ | $3.40\times10^{3}$ |
| | 1 min | $4.64\times10^{9}$ | 4 d p.i. | $8.23\times10^{7}$ | $1.20\times10^{4}$ |
| | 5 min | $3.66\times10^{9}$ | 4 d p.i. | $1.19\times10^{8}$ | $9.20\times10^{3}$ |
| | 15 min | $2.93\times10^{9}$ | 4 d p.i. | $4.50\times10^{7}$ | $6.27\times10^{2}$ |
| | 30 min | $4.31\times10^{9}$ | 4 d p.i. | $1.65\times10^{8}$ | $1.58\times10^{4}$ |
| | 45 min | $4.07\times10^{9}$ | 4 d p.i. | $8.91\times10^{7}$ | $3.15\times10^{3}$ |
| | 60 min | $4.30\times10^{9}$ | 4 d p.i. | $1.01\times10^{8}$ | $2.06\times10^{4}$ |
| | 90 min | $1.83\times10^{9}$ | 4 d p.i. | $1.85\times10^{8}$ | $1.20\times10^{4}$ |
| | 120 min | $1.67\times10^{9}$ | 4 d p.i. | $6.51\times10^{7}$ | $4.12\times10^{3}$ |
| | 240 min | $3.12\times10^{4}$ | 4 d p.i. | $4.61\times10^{1}$ | $5.59\times10^{1}$* |
| P2 | Swab | $1.70\times10^{9}$ | 10 d p.i. | $3.24\times10^{7}$ | $3.40\times10^{2}$ |
| | 1 min | $1.27\times10^{9}$ | 10 d p.i. | $8.94\times10^{7}$ | $5.59\times10^{1}$* |
| | 5 min | $5.63\times10^{0}$ | no | n.d. | $5.59\times10^{1}$* |
| | 15 min | $5.49\times10^{8}$ | 10 d p.i. | $7.45\times10^{7}$ | $5.59\times10^{1}$* |
| | 30 min | $6.40\times10^{1}$ | no | n.d. | $5.59\times10^{1}$* |
| | 45 min | $2.55\times10^{2}$ | no | n.d. | $5.59\times10^{1}$* |
| | 60 min | n.d. | no | n.d. | $5.59\times10^{1}$* |
| | 90 min | $3.73\times10^{0}$ | no | n.d. | $5.59\times10^{1}$* |
| | 120 min | n.d. | no | n.d. | $5.59\times10^{1}$* |
| | 240 min | $5.01\times10^{2}$ | no | $2.19\times10^{1}$ | $5.59\times10^{1}$* |
| P3 | Swab | $1.35\times10^{9}$ | 3 d p.i. | $5.31\times10^{7}$ | $2.32\times10^{4}$ |
| | 1 min | $1.27\times10^{9}$ | 3 d p.i. | $1.64\times10^{8}$ | $5.90\times10^{4}$ |
| | 5 min | $3.15\times10^{9}$ | 3 d p.i. | $1.38\times10^{8}$ | $2.26\times10^{5}$ |
| | 15 min | $3.21\times10^{9}$ | 3 d p.i. | $1.52\times10^{8}$ | $2.26\times10^{5}$ |
| | 30 min | $5.47\times10^{9}$ | 3 d p.i. | $1.73\times10^{8}$ | $3.86\times10^{5}$ |
| | 45 min | $1.89\times10^{9}$ | 3 d p.i. | $1.21\times10^{8}$ | $3.87\times10^{5}$ |
| | 60 min | $3.59\times10^{9}$ | 3 d p.i. | $1.39\times10^{8}$ | $2.96\times10^{5}$ |
| | 90 min | $4.09\times10^{9}$ | 3 d p.i. | $1.81\times10^{8}$ | $8.68\times10^{5}$ |
| | 120 min | $5.90\times10^{9}$ | 3 d p.i. | $1.56\times10^{8}$ | $6.62\times10^{5}$ |
| | 240 min | $2.68\times10^{9}$ | 3 d p.i. | $1.65\times10^{8}$ | $1.13\times10^{6}$ |
| P4 | Swab | Excluded due to bacterial/fungal contamination | | | |
| | 1 min | | | | |
| | 5 min | | | | |
| | 15 min | | | | |
| | 30 min | | | | |
| | 45 min | | | | |
| | 60 min | | | | |
| | 90 min | | | | |
| | 120 min | | | | |
| | 240 min | | | | |

| | | | | | |
|---|---|---|---|---|---|
| P5 | Swab | $1.55\times10^{8}$ | 10 d p.i. | $1.99\times10^{7}$ | $1.58\times10^{2}$ |
| | 1 min | $5.49\times10^{8}$ | 10 d p.i. | $1.40\times10^{8}$ | $7.73\times10^{4}$ |
| | 5 min | $5.20\times10^{8}$ | 10 d p.i. | $1.47\times10^{8}$ | $1.58\times10^{4}$ |
| | 15 min | $1.70\times10^{8}$ | 10 d p.i. | $1.39\times10^{8}$ | $1.58\times10^{4}$ |
| | 30 min | $3.94\times10^{8}$ | 10 d p.i. | $1.80\times10^{8}$ | $3.53\times10^{4}$ |
| | 45 min | $4.30\times10^{8}$ | 10 d p.i. | $1.69\times10^{8}$ | $7.73\times10^{4}$ |
| | 60 min | $2.39\times10^{8}$ | 10 d p.i. | $1.03\times10^{8}$ | $4.80\times10^{2}$ |
| | 90 min | $4.33\times10^{8}$ | 10 d p.i. | $2.34\times10^{8}$ | $9.20\times10^{3}$ |
| | 120 min | $3.79\times10^{8}$ | 10 d p.i. | $1.08\times10^{8}$ | $2.06\times10^{4}$ |
| | 240 min | $3.13\times10^{8}$ | 10 d p.i. | $1.35\times10^{8}$ | $2.06\times10^{4}$ |
| P6 | Swab | $1.22\times10^{8}$ | 10 d p.i. | $2.26\times10^{7}$ | $5.59\times10^{1}$* |
| | 1 min | $2.71\times10^{8}$ | no | $8.74\times10^{7}$ | $2.95\times10^{5}$ |
| | 5 min | $2.86\times10^{2}$ | no | $5.88\times10^{1}$ | $5.59\times10^{1}$* |
| | 15 min | $6.66\times10^{7}$ | no | $1.11\times10^{8}$ | $2.40\times10^{3}$ |
| | 30 min | $3.80\times10^{2}$ | no | $3.36\times10^{1}$ | $5.59\times10^{1}$* |
| | 45 min | $2.40\times10^{3}$ | no | $2.30\times10^{1}$ | $5.59\times10^{1}$* |
| | 60 min | $6.08\times10^{0}$ | no | n.d. | $5.59\times10^{1}$* |
| | 90 min | $2.70\times10^{1}$ | no | $4.63\times10^{2}$ | $5.59\times10^{1}$* |
| | 120 min | n.d. | no | $1.11\times10^{1}$ | $5.59\times10^{1}$* |
| | 240 min | $1.18\times10^{1}$ | no | $2.76\times10^{1}$ | $5.59\times10^{1}$* |
| P7 | Swab | $5.62\times10^{7}$ | no | $3.82\times10^{7}$ | $5.59\times10^{1}$* |
| | 1 min | n.d. | no | n.d. | $5.59\times10^{1}$* |
| | 5 min | n.d. | no | n.d. | $5.59\times10^{1}$* |
| | 15 min | $1.09\times10^{1}$ | no | $1.49\times10^{1}$ | $5.59\times10^{1}$* |
| | 30 min | n.d. | no | n.d. | $5.59\times10^{1}$* |
| | 45 min | n.d. | no | n.d. | $5.59\times10^{1}$* |
| | 60 min | $6.90\times10^{0}$ | no | n.d. | $5.59\times10^{1}$* |
| | 90 min | $4.95\times10^{0}$ | no | n.d. | $5.59\times10^{1}$* |
| | 120 min | n.d. | no | $4.36\times10^{7}$ | $5.59\times10^{1}$* |
| | 240 min | $6.15\times10^{0}$ | no | $9.37\times10^{6}$ | $5.59\times10^{1}$* |
| P8 | Swab | $6.78\times10^{8}$ | 10 d p.i. | $4.51\times10^{7}$ | $1.58\times10^{4}$ |
| | 1 min | $8.54\times10^{6}$ | no | $8.09\times10^{6}$ | $5.59\times10^{1}$* |
| | 5 min | n.d. | no | n.d. | $5.59\times10^{1}$* |
| | 15 min | $2.52\times10^{1}$ | no | n.d. | $5.59\times10^{1}$* |
| | 30 min | n.d. | no | n.d. | $5.59\times10^{1}$* |
| | 45 min | n.d. | no | n.d. | $5.59\times10^{1}$* |
| | 60 min | $1.02\times10^{2}$ | no | n.d. | $5.59\times10^{1}$* |
| | 90 min | n.d. | no | n.d. | $5.59\times10^{1}$* |
| | 120 min | n.d. | no | n.d. | $5.59\times10^{1}$* |
| | 240 min | n.d. | no | n.d. | $5.59\times10^{1}$* |
| P9 | Swab | $2.03\times10^{9}$ | 10 d p.i. | $2.94\times10^{7}$ | $5.59\times10^{1}$* |
| | 1 min | n.d. | no | n.d. | $5.59\times10^{1}$* |
| | 5 min | n.d. | no | n.d. | $5.59\times10^{1}$* |
| | 15 min | $2.09\times10^{9}$ | 10 d p.i. | n.d. | $5.59\times10^{1}$* |
| | 30 min | $3.33\times10^{0}$ | no | n.d. | $5.59\times10^{1}$* |
| | 45 min | $8.39\times10^{1}$ | no | n.d. | $5.59\times10^{1}$* |
| | 60 min | n.d. | no | n.d. | $5.59\times10^{1}$* |

| | | | | | |
|---|---|---|---|---|---|
| | 90 min | n.d. | no | n.d. | $5.59\times10^{1}$* |
| | 120 min | $3.07\times10^{0}$ | no | n.d. | $5.59\times10^{1}$* |
| | 240 min | $1.03\times10^{1}$ | no | n.d. | $5.59\times10^{1}$* |
| P10 | Swab | $2.00\times10^{8}$ | 10 d p.i. | $6.58\times10^{7}$ | $2.06\times10^{4}$ |
| | 1 min | $1.86\times10^{9}$ | 10 d p.i. | $2.85\times10^{8}$ | $3.53\times10^{4}$ |
| | 5 min | n.d. | no | $3.64\times10^{1}$ | $5.59\times10^{1}$* |
| | 15 min | $3.86\times10^{1}$ | no | $5.82\times10^{1}$ | $5.59\times10^{1}$* |
| | 30 min | $3.06\times10^{1}$ | no | $4.83\times10^{1}$ | $1.57\times10^{4}$ |
| | 45 min | $8.76\times10^{1}$ | no | $8.42\times10^{1}$ | $9.22\times10^{3}$ |
| | 60 min | $6.13\times10^{1}$ | no | $4.55\times10^{1}$ | $5.59\times10^{1}$* |
| | 90 min | $9.36\times10^{0}$ | no | $4.22\times10^{1}$ | $5.59\times10^{1}$* |
| | 120 min | $1.98\times10^{0}$ | no | $4.42\times10^{1}$ | $5.59\times10^{1}$* |
| | 240 min | $1.38\times10^{1}$ | no | $2.54\times10^{2}$ | $5.59\times10^{1}$* |
| P11 | Swab | $2.00\times10^{8}$ | 10 d p.i. | $1.72\times10^{6}$ | $5.59\times10^{1}$* |
| | 1 min | $1.67\times10^{0}$ | no | n.d. | $5.59\times10^{1}$* |
| | 5 min | $7.71\times10^{0}$ | no | n.d. | $5.59\times10^{1}$* |
| | 15 min | n.d. | no | n.d. | $5.59\times10^{1}$* |
| | 30 min | n.d. | no | n.d. | $5.59\times10^{1}$* |
| | 45 min | n.d. | no | n.d. | $5.59\times10^{1}$* |
| | 60 min | n.d. | no | n.d. | $5.59\times10^{1}$* |
| | 90 min | n.d. | no | n.d. | $5.59\times10^{1}$* |
| | 120 min | n.d. | no | n.d. | $5.59\times10^{1}$* |
| | 240 min | n.d. | no | n.d. | $5.59\times10^{1}$* |
| P12 | Swab | $2.23\times10^{1}$ | no | n.d. | $5.59\times10^{1}$* |
| | 1 min | n.d. | no | n.d. | $5.59\times10^{1}$* |
| | 5 min | n.d. | no | n.d. | $5.59\times10^{1}$* |
| | 15 min | n.d. | no | n.d. | $5.59\times10^{1}$* |
| | 30 min | n.d. | no | n.d. | $5.59\times10^{1}$* |
| | 45 min | n.d. | no | n.d. | $5.59\times10^{1}$* |
| | 60 min | n.d. | no | n.d. | $5.59\times10^{1}$* |
| | 90 min | $4.00\times10^{6}$ | no | n.d. | $5.59\times10^{1}$* |
| | 120 min | n.d. | no | n.d. | $5.59\times10^{1}$* |
| | 240 min | n.d. | no | n.d. | $5.59\times10^{1}$* |
| P13 | Swab | $2.75\times10^{1}$ | no | n.d. | $5.59\times10^{1}$* |
| | 1 min | n.d. | no | n.d. | $5.59\times10^{1}$* |
| | 5 min | n.d. | no | n.d. | $5.59\times10^{1}$* |
| | 15 min | n.d. | no | n.d. | $5.59\times10^{1}$* |
| | 30 min | n.d. | no | n.d. | $5.59\times10^{1}$* |
| | 45 min | n.d. | no | n.d. | $5.59\times10^{1}$* |
| | 60 min | n.d. | no | n.d. | $5.59\times10^{1}$* |
| | 90 min | n.d. | no | n.d. | $5.59\times10^{1}$* |
| | 120 min | n.d. | no | n.d. | $5.59\times10^{1}$* |
| | 240 min | n.d. | no | n.d. | $5.59\times10^{1}$* |
| P14 | Swab | $3.28\times10^{4}$ | no | $1.35\times10^{4}$ | $5.59\times10^{1}$* |
| | 1 min | n.d. | no | n.d. | $5.59\times10^{1}$* |
| | 5 min | n.d. | no | n.d. | $5.59\times10^{1}$* |

| | | | | | |
|---|---|---|---|---|---|
| | 15 min | n.d. | no | n.d. | $5.59\times10^{1}$* |
| | 30 min | n.d. | no | n.d. | $5.59\times10^{1}$* |
| | 45 min | n.d. | no | n.d. | $5.59\times10^{1}$* |
| | 60 min | n.d. | no | n.d. | $5.59\times10^{1}$* |
| | 90 min | n.d. | no | n.d. | $5.59\times10^{1}$* |
| | 120 min | n.d. | no | n.d. | $5.59\times10^{1}$* |
| | 240 min | n.d. | no | n.d. | $5.59\times10^{1}$* |
| P15 | Swab | | | | |
| | 1 min | | | | |
| | 5 min | | | | |
| | 15 min | | | | |
| | 30 min | Excluded due to bacterial/fungal contamination | | | |
| | 45 min | | | | |
| | 60 min | | | | |
| | 90 min | | | | |
| | 120 min | | | | |
| | 240 min | | | | |

* lower limit of quantification (LLOQ); n.d. not detectable

**Appendix figures**

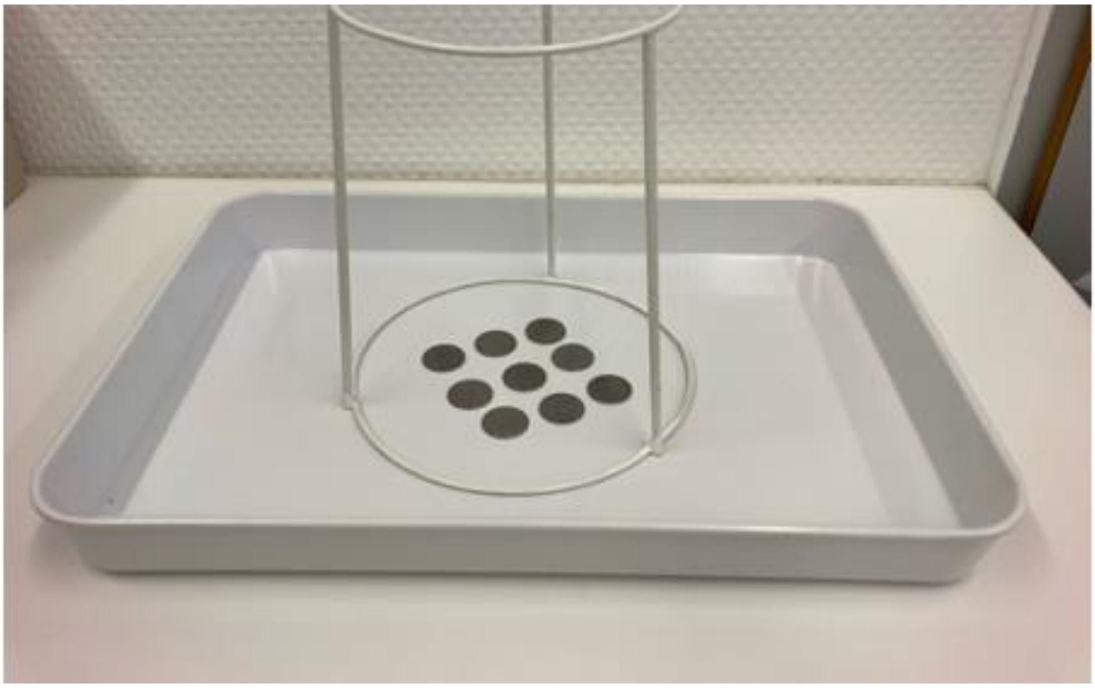

**Appendix Figure 1: Experimental setup for contaminating samples with coughing.**

Nine steel-carriers (each with a diameter of one centimeter) were placed under a specially designed tripod with a defined distance of 15 centimeters. Patients were asked to rest their face in the opening of the tripod and were encouraged to forcefully cough on the pre-defined surface area twice. After defined time points at room temperature (1 min, 5 min, 15 min, 30 min, 45 min, 90 min, 120 min and 240 min), the steel-carriers were placed in cell culture medium and virus rescue was attempted in cell culture.

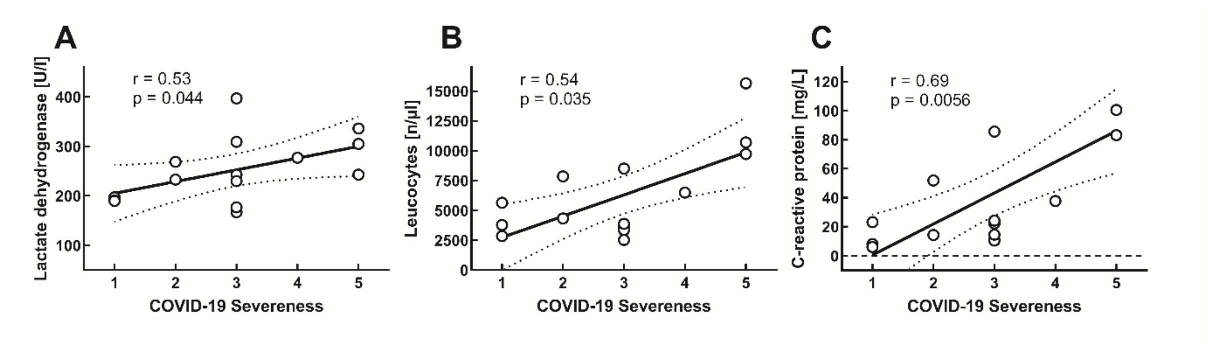


**Appendix Figure 2: Correlation of laboratory findings and COVID-19 severeness.**

Correlations of COVID-19 severeness with lactate dehydrogenase (panel A), leucocytes (panel B) and C-reactive protein (panel C). COVID-19 severeness was rated according to WHO-scoring ) [9] from zero to five (0 = no apparent symptoms of COVID-19, 1 = mild, 2 = moderate, 3 = severe, 4 = critical severeness, 5 = death). Correlation was calculated using Spearman’s correlation coefficient.

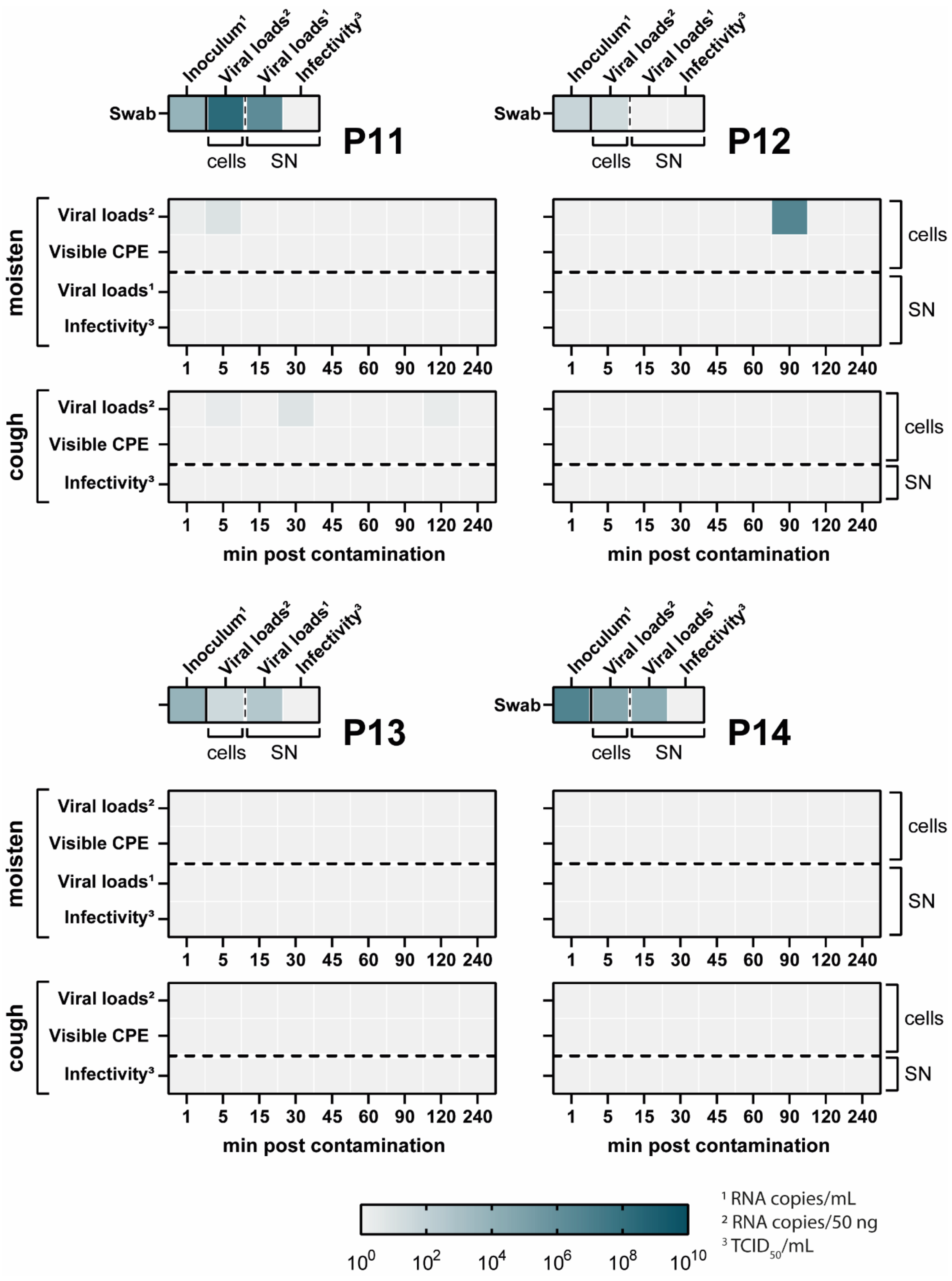


**Appendix Figure 3: Quantification of viral loads and infectivity of patient swabs, "moisten" samples and "cough" samples that could not be successfully recovered in cell culture.**

VeroE6 cells were inoculated with the patient material and monitored daily. Upon the emergence of cytopathic effects, the supernatant was collected to determine viral loads by RT-qPCR (RNA copies/mL; indicated by [1]) and viral titers by an endpoint-dilution assay ($TCID_{50}$/mL, indicated by [3]). RNA was isolated from the cells and subjected to RT-qPCR to determine viral loads (RNA copies/50 ng total RNA, indicated by [2]). For each patient (P11-P14), three panels were designed. The top small panel includes data of the patient swabs alone, while the larger middle panel shows the data for the "moistened" samples and the lower panel the data from the "cough" samples. For "moisten" and "cough" samples viral loads and infectivity at nine different time points were determined. The color indicates the amount of virus being detectable in each sample, with light grey being the lower limit of detection to dark green resulting in $10^{10}$ RNA copies/mL, RNA copies/50 ng or $TCID_{50}$/mL. The visible CPE was rated two dimensionally, with light grey being "no visible CPE" and dark green being "visible CPE".
